# Familial Risk and Heritability of Lower Urinary Tract Symptoms in Children and Young Adults

**DOI:** 10.1101/2025.03.15.25324031

**Authors:** Lotta Renström-Koskela, Viktor Skokic, Danielle Scharp, Joseph D. Buxbaum, Dorothy E. Grice, Behrang Mahjani, Olof Akre

## Abstract

**Importance:** Lower urinary tract symptoms (LUTS) in children and young adults are common and can significantly impact quality of life. While prior studies suggest a genetic component, the extent of familial aggregation and heritability remains unclear.

**Objective:** To estimate the familial aggregation and heritability of LUTS in a nationwide cohort of children and young adults.

**Design, Setting, and Participants:** This population-based cohort study used data from Swedish national registers to identify all live-born singleton children in Sweden between January 1, 1995, and December 31, 2005, with follow-up through December 31, 2018. Familial relationships were determined using the Swedish Multi-Generation Register, and LUTS cases were identified using ICD-10 diagnoses and prescription data from the Swedish National Patient Register and Swedish Prescribed Drug Register.

**Exposure:** Family history of LUTS, defined by sibling and cousin relationships.

**Main Outcomes and Measures:** Familial aggregation of LUTS was assessed by estimating relative recurrence risks (RRs) with 95% CIs among siblings and cousins of affected individuals. Heritability was estimated using Falconer’s Liability Threshold Model.

**Results:** Among 982,903 individuals, LUTS cases were identified using ICD-10, narrow ICD-medication, and broad ICD-medication definitions. Familial aggregation was strongest among siblings, with relative risks of 7.41 (95% CI, 5.72–9.60; P < .001) for storage LUTS and 6.53 (95% CI, 2.09–20.38; P = .001) for voiding LUTS using the narrow ICD-medication definition. Cousin recurrence risks were lower, supporting a genetic contribution to LUTS. Sex-specific analyses showed higher familial aggregation in same-sex sibling pairs, particularly for voiding LUTS in female-female pairs (RR, 14.22; 95% CI, 1.97–102.82; P = .009).

Heritability estimates further supported a genetic basis for LUTS. Storage LUTS heritability was 55% (95% CI, 45.5%–64.7%) using the narrow ICD-medication definition and 48% (95% CI, 46.8%–48.6%) using the broad definition. Voiding LUTS heritability was lower, at 40% (95% CI, 37.6%–43.2%) and 26% (95% CI, 14.8%–37.7%) for the narrow and broad definitions, respectively.

**Conclusions and Relevance:** These findings provide strong evidence for a genetic contribution to LUTS in children and young adults, with higher familial aggregation among closely related individuals and same-sex sibling pairs. The moderate to high heritability estimates suggest that genetic factors play a key role in LUTS susceptibility, with potential sex-specific influences. These findings may inform risk stratification, early intervention strategies, and future genetic research.

**Key Points:** *Question:* To what extent do genetic factors contribute to lower urinary tract symptoms (LUTS) in children and young adults?

*Findings:* In this population-based cohort study of 982,903 individuals, LUTS exhibited strong familial aggregation, with siblings of affected individuals having up to a 7.41-fold increased risk. Heritability estimates ranged from 26% to 55%, suggesting a substantial genetic contribution.

*Meaning:* These findings support a strong genetic basis for LUTS, with potential sex-specific genetic influences, highlighting the need for risk prediction and genetic research in LUTS.

## 1. Introduction

Lower urinary tract symptoms (LUTS), including urgency, frequency, nocturia, and urinary incontinence, are common conditions that can significantly affect physical health, psychological well-being, and social functioning. Although LUTS have been widely studied in older adults, growing evidence highlights their substantial prevalence in younger populations, affecting up to 20% of children and adolescents, and their potential long-term consequences (1–3).^1–3^ In children, adolescents, and young adults, LUTS can interfere with daily activities, academic performance, and social development, contributing to emotional distress and reduced self-esteem.^4,5^ Moreover, symptoms that begin in childhood often persist into adulthood, increasing the risk of chronic urological disorders, anxiety, depression, and diminished quality of life.^6,7^ Despite these concerns, LUTS in younger individuals remain underrecognized and understudied, particularly with regard to their genetic and familial determinants.

There is increasing evidence that genetic factors play a key role in LUTS susceptibility. Familial clustering has been observed in conditions such as nocturnal enuresis, where first-degree relatives have a markedly higher risk than the general population.^8,9^ Twin studies further support a genetic contribution, with estimates suggesting that 30–60% of the variance in overactive bladder syndrome is attributable to genetic factors.^10^ Additionally, emerging genome-wide association studies (GWAS) have identified genetic loci potentially involved in bladder dysfunction, suggesting underlying neurogenic and myogenic mechanisms; however, these findings remain preliminary and require further validation.^11^ Despite these insights, most existing research is limited by small sample sizes, inconsistent diagnostic criteria, and a predominant focus on adult populations. As a result, the familial and genetic contributions to LUTS in children and young adults remain poorly understood, highlighting the need for large-scale, population-based studies to address these gaps.

Using Swedish national register data, our previous research identified age- and sex-specific variations in LUTS incidence, suggesting a potential genetic influence.^12^ The comprehensive and longitudinal nature of these registers, along with detailed familial linkage information, provides a unique opportunity to investigate familial aggregation and estimate heritability in a large, population-based cohort.^13^ In this study, we aim to (1) quantify familial risk of LUTS across different age groups and sexes, (2) estimate heritability using validated epidemiological models, and (3) examine sex-specific familial transmission patterns.

Understanding the genetic and familial contributions to LUTS has important clinical and public health implications. Identifying individuals at higher genetic risk could facilitate earlier diagnosis, improved risk stratification, and personalized treatment strategies. Furthermore, elucidating the genetic architecture of LUTS may provide insights into underlying biological mechanisms—such as neurotransmitter signaling, inflammatory pathways, or structural abnormalities—that could inform the development of targeted therapies and advance precision medicine approaches.^14,15^ By integrating epidemiological and genetic data, this study aims to enhance understanding of LUTS etiology, ultimately leading to improved patient outcomes and more effective clinical management strategies.

## 2. Methods

### 2.1 Study Design and Population

This population-based cohort study used data from Swedish national registers to investigate familial aggregation and heritability of LUTS. The study included all live-born singleton children in Sweden between January 1, 1995, and December 31, 2005, as recorded in the Swedish Medical Birth Register, with follow-up through December 31, 2018, or until emigration. After excluding 30,840 non-singleton births and one individual with missing sex data, the final cohort comprised 982,903 individuals.

The Swedish healthcare system provides universal coverage, with health and demographic data recorded in national registers. The Swedish Medical Birth Register, which has captured approximately 98% of all births in Sweden since 1973, and the Swedish Multi-Generation Register, which links individuals to their biological and adoptive parents, enabled identification of familial relationships and disease outcomes. These registers have been validated for epidemiologic research.^16–18^

Familial relationships were established using the Swedish Multi-Generation Register, which links individuals born in Sweden from 1932 onward to their biological parents. Degrees of genetic relatedness were categorized into first-degree, second-degree, and third-degree relationships. First-degree relatives included parent-offspring pairs and full siblings, who share approximately 50% of their genetic material. Second-degree relatives included half-siblings, grandparent-grandchild pairs, and avuncular (aunt/uncle-niece/nephew) relationships, corresponding to approximately 25% shared genetic material. Third-degree relatives included first cousins, who share approximately 12.5% of their genetic material. This classification enabled the assessment of familial aggregation at varying levels of genetic relatedness while controlling for shared environmental factors.

### 2.2 LUTS Case Identification

LUTS cases were identified using three predefined criteria.^12^ First, LUTS was identified using ICD-10 diagnostic codes in the Swedish National Patient Register. Storage symptoms were defined using ICD-10 codes N39.4A, N39.4C, and R35.9B, while voiding symptoms were defined using R33.9. Individuals with neurogenic bladder were excluded.^19^ Second, a broad ICD-medication definition was applied, in which cases were classified if they had an ICD-10 diagnosis or at least one prescription of LUTS-specific medications recorded in the Swedish Prescribed Drug Register. These medications included alpha-1 receptor inhibitors for voiding symptoms and anticholinergics or beta-3 receptor agonists for storage symptoms. Third, a narrow ICD-medication definition was applied, requiring an ICD-10 diagnosis or at least two prescriptions of LUTS-specific medications dispensed more than 76 days apart within one year. Medication-based definitions were included to capture cases managed in primary care, which are underrepresented in hospital-based registers.

### 2.3 Statistical Analysis

Cox proportional hazards regression models were used to estimate hazard ratios (HRs) with 95% confidence intervals (CIs), comparing the risk of LUTS among relatives of affected versus unaffected individuals. For each familial relationship (siblings, half-siblings, and cousins), all possible relative pairs were identified. Using one individual’s LUTS status as the predictor (index case) and the relative’s status as the outcome, HRs were estimated to quantify relative recurrence risk.

Follow-up time was calculated from birth until LUTS diagnosis, emigration, death, or December 31, 2018. Models were adjusted for birth year, sex, parental age at birth, and socioeconomic status. A robust sandwich estimator accounted for non-independence within sibling pairs. Separate analyses were conducted for storage and voiding LUTS using all three diagnostic definitions. A sibling and cousins comparison design was used to account for unmeasured familial confounders, such as genetic predisposition and early-life environmental exposures.

Sex-stratified analyses were conducted to assess differences in familial aggregation patterns between male and female relatives. Same-sex pairs (male-male, female-female) and opposite-sex pairs (male-female) were analyzed separately.

Heritability was estimated using Falconer’s Liability Threshold Model, which quantifies genetic contributions to disease risk while accounting for shared environmental factors.^20,21^ Familial concordance data were used to estimate variance components, and weighted least squares regression was applied to compute heritability estimates for storage and voiding LUTS.

The study was approved by the Regional Ethical Review Board in Stockholm, Sweden. A waiver of informed consent was granted, as individual consent is not required for national register-based research in Sweden.

## 3. Results

Among the 982,903 individuals in the study cohort, LUTS cases were identified using three predefined criteria: ICD-10 diagnosis, broad ICD-medication definition, and narrow ICD-medication definition. As previously reported, the ICD-10 definition identified 633 individuals (0.1%) with storage LUTS and 1,116 individuals (0.1%) with voiding LUTS (Table 1). Using the broad ICD-medication definition, 8,825 individuals (0.9%) were classified as having storage LUTS and 1,998 individuals (0.2%) as having voiding LUTS. The narrow ICD-medication definition identified 4,894 individuals (0.5%) with storage LUTS and 1,215 individuals (0.1%) with voiding LUTS.

**Table 1.** Baseline Characteristics of the Study Cohort.

|  | Total Population |  | Females |  |
| --- | --- | --- | --- | --- |
|  | Without LUTS<br>n (%) | With LUTS<br>n (%) | Without LUTS<br>n (%) | With LUTS<br>n (%) |
| Storage LUTS<br>ICD-10 | 982,270 (99.9) | 633 (0.1) | 477,081 (99.9) | 359 (0.1) |
| Storage LUTS<br>Narrow ICD-Medication | 974,078 (99.1) | 8,825 (0.9) | 473,447 (99.2) | 3,993 (0.8) |
| Storage LUTS<br>Broad ICD-medication | 978,009 (99.5) | 4,894 (0.5) | 475,376 (99.6) | 2,064 (0.4) |
| Voiding LUTS<br>ICD-10 | 981,787 (99.9) | 1,116 (0.1) | 476,971 (99.9) | 469 (0.1) |
| Voiding LUTS<br>Narrow ICD-Medication | 980,905 (99.8) | 1,998 (0.2) | 476,541 (99.8) | 899 (0.2) |
| Voiding LUTS<br>Broad ICD-Medication | 980,905 (99.8) | 1,215 (0.1) | 476,929 (99.9) | 511 (0.1) |

Familial aggregation analyses demonstrated significantly increased risk among siblings of individuals with LUTS compared to siblings of unaffected individuals (Table 2). For storage LUTS using the narrow ICD-medication definition, siblings had a relative recurrence risk (RR) of 7.41 (95% CI, 5.72–9.60; P < .001). With the broader definition, the RR remained elevated at 4.96 (95% CI, 4.16–5.90; P < .001). Voiding LUTS also demonstrated significant sibling aggregation, with an RR of 6.53 (95% CI, 2.09–20.38; P = .001) for the narrow ICD-medication definition and 3.23 (95% CI, 1.21–8.61; P = .019) for the broad ICD-medication definition.

**Table 2.**
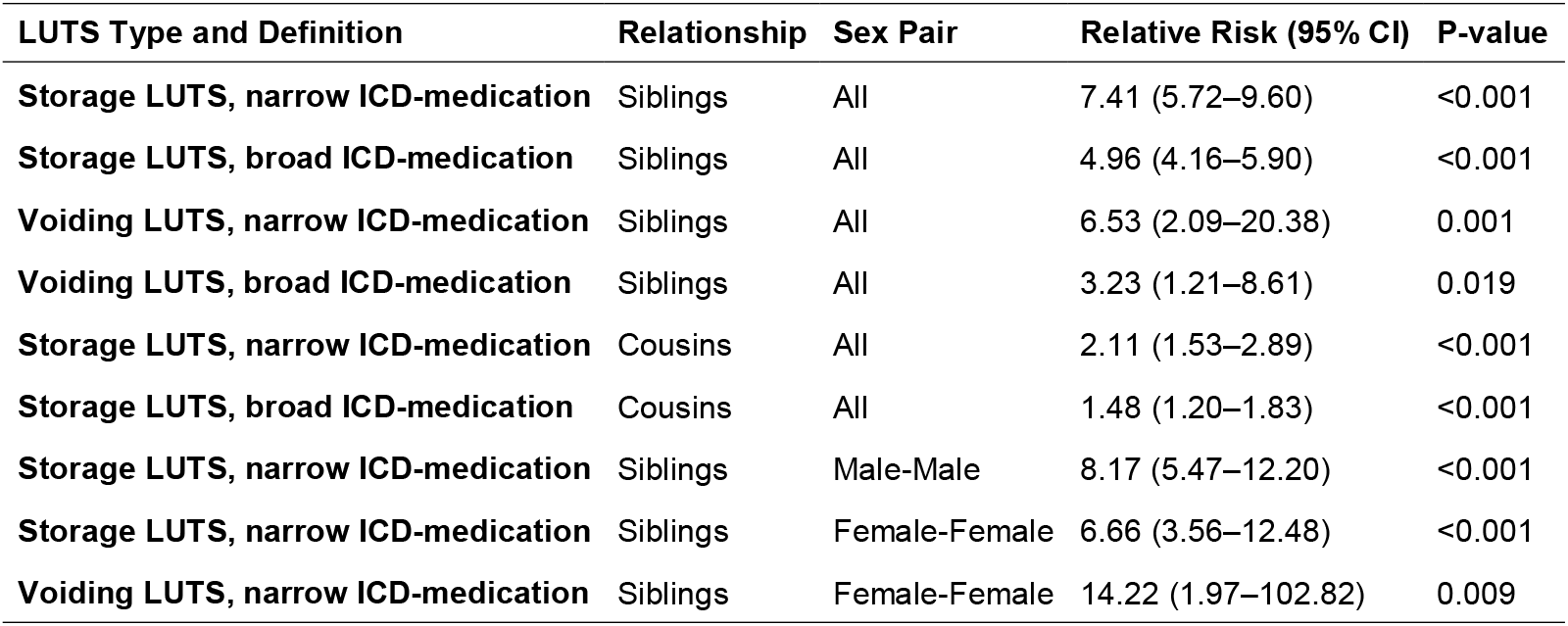
Familial Aggregation and Sex-specific Risks for LUTS.

Familial aggregation decreased as genetic relatedness became more distant. Cousins of individuals with storage LUTS had lower but still significantly elevated risks, with an RR of 2.11 (95% CI, 1.53–2.89; P < .001) using the narrow definition and 1.48 (95% CI, 1.20–1.83; P < .001) using the broader definition. For voiding LUTS, cousin recurrence risks were not statistically significant.

Sex-specific analyses uncovered notable differences in familial aggregation patterns. For storage LUTS, male-male sibling pairs exhibited the highest RR at 8.17 (95% CI, 5.47–12.20; P < .001), while female-female sibling pairs had slightly lower but still significantly elevated risk at 6.66 (95% CI, 3.56–12.48; P < .001). Conversely, for voiding LUTS, female-female sibling pairs demonstrated a substantially higher risk, with an RR of 14.22 (95% CI, 1.97–102.82; P = .009). Data for male-male sibling pairs in voiding LUTS was insufficient for analysis.

Detailed subgroup analyses by birth year indicated robust findings across different birth cohorts, with few exceptions. Notably, for storage LUTS (narrow ICD-medication), slightly elevated recurrence risks were observed in specific birth years: RR of 1.17 (95% CI, 1.00–1.36; P = .052) in 2002, and RR of 1.20 (95% CI, 1.03–1.40; P = .023) in 2003.

Heritability analyses using Falconer’s Liability Threshold Model strongly supported a genetic component to LUTS susceptibility (Table 3). Storage LUTS heritability estimates were 55% (95% CI, 45.5%–64.7%) using the narrow ICD-medication definition and 48% (95% CI, 46.8%–48.6%) using the broad definition. Voiding LUTS had lower heritability estimates, at 40% (95% CI, 37.6%–43.2%) and 26% (95% CI, 14.8%–37.7%) for the narrow and broad definitions, respectively.

**Table 3.** Heritability Estimates (h^2^) for LUTS by Type and Definition.

| LUTS Type | Heritability ($h^2$) [95% CI] |
| --- | --- |
| Storage LUTS - Narrow ICD-Med | 0.551 (0.455–0.647) |
| Storage LUTS - Broad ICD-Med | 0.477 (0.468–0.486) |
| Voiding LUTS - Narrow ICD-Med | 0.404 (0.376–0.432) |
| Voiding LUTS - Broad ICD-Med | 0.262 (0.148–0.377) |

## 4. Discussion

This nationwide, population-based study provides robust evidence for a substantial genetic contribution to LUTS in children and young adults. Using comprehensive Swedish national registers, we identified significant familial aggregation of LUTS, clearly demonstrating that relative risks decline as genetic relatedness decreases. Siblings of affected individuals had substantially elevated risks—up to 7.41-fold for storage LUTS and 6.53-fold for voiding LUTS compared to siblings of unaffected individuals. Recurrence risks among cousins, while lower, remained significantly elevated for storage LUTS, further emphasizing genetic contributions.

Our findings regarding familial aggregation align closely with previous studies exploring familial clustering in related urological conditions, such as nocturnal enuresis, where recurrence risks among first-degree relatives were reported between 5-to 7-fold increased compared to the general population, suggesting potential shared genetic mechanisms underlying these conditions.^22,23^

Sex-specific analyses revealed distinct familial aggregation patterns, indicating potential sex-specific genetic susceptibility to LUTS. Male-male sibling pairs showed particularly strong aggregation for storage LUTS, whereas female-female sibling pairs demonstrated notably high aggregation for voiding LUTS. Although wide confidence intervals in female-female pairs suggest limited statistical power due to fewer cases, these preliminary observations highlight the importance of further investigations into sex-specific genetic mechanisms. Prior studies support this hypothesis, reporting substantial sex differences in LUTS prevalence and clinical manifestation.^24,25^ Future research utilizing larger, specifically designed cohorts could better elucidate these sex-specific genetic factors.

Heritability analyses using Falconer’s Liability Threshold Model provided additional strong support for genetic contributions to LUTS susceptibility. Storage LUTS heritability estimates were notably higher (55% narrow definition, 48% broad definition) compared to voiding LUTS (40% narrow definition, 26% broad definition), suggesting environmental factors might play a larger role in voiding symptoms. This aligns with previous twin studies that reported similar heritability estimates (30%-60%) for related urinary conditions, further validating our findings and suggesting that storage symptoms may have stronger underlying genetic components compared to voiding symptoms.^26,27^ The lower heritability of voiding LUTS points to potentially greater environmental contributions, which could include early-life toilet-training practices, diet, psychological stressors, or infections.

The present study have several key strengths. The large, population-based cohort and comprehensive follow-up through validated national registers enabled robust, generalizable conclusions. The use of multiple case definitions enhanced diagnostic validity across diverse healthcare settings, and detailed familial relationship analyses offered novel insights into the genetic architecture of LUTS.

However, several limitations warrant consideration. The reliance on register-based diagnoses likely led to underestimating milder LUTS cases, as individuals managed with lifestyle modifications or over-the-counter treatments were not captured. The study design also did not allow for detailed assessment of symptom severity or specific LUTS subtypes, which might differ genetically. Moreover, younger individuals in the cohort had fewer years of observation, potentially affecting diagnostic ascertainment rates. Future research incorporating clinical evaluations and patient questionnaires could clarify symptom severity and subtype differences, thereby refining understanding of genetic and environmental contributions.

These findings have significant implications for clinical practice and future research. Clinicians should actively consider family history when assessing LUTS risk, facilitating earlier diagnosis and intervention among high-risk individuals. The moderate to high heritability of LUTS underscores the importance of genetic research to identify genetic variants that could enhance risk prediction, early intervention, and targeted therapies. The observed sex-specific genetic patterns emphasize the need for personalized diagnostic and therapeutic approaches, tailored to patient sex and specific LUTS subtype.

In conclusion, this large-scale, population-based study provides compelling evidence for a significant genetic basis underlying LUTS in children and young adults. Pronounced familial aggregation, clear sex-specific differences, and substantial heritability estimates underscore the importance of genetic factors in LUTS susceptibility, providing essential groundwork for future research, risk stratification, and the development of targeted interventions aimed at improving patient outcomes.

## Data Availability

Data from the Swedish national registers may be obtained from a third party and are not publicly available. Data cannot be shared publicly owing to restrictions by law. Data are available from the National Medical Registries in Sweden after approval by the Swedish Ethical Review Authority.

## Acknowledgments

This study was supported by a grant from the Swedish Research Council (2021-02846) and the Beatrice and Samuel A. Seaver Foundation, Icahn School of Medicine at Mount Sinai, New York, NY.

## Author Contributions

<u>Study concepts and design:</u> OA, JDB, DG, BM, LRK, VS

<u>Acquisition, analysis, or interpretation of data:</u> OA, JDB, DG, BM, LRK, VS

<u>Drafting of the manuscript:</u> All authors

<u>Critical revision of the manuscript for important intellectual content:</u> All authors

<u>Statistical analysis:</u> BM, VS

<u>Obtained funding:</u> OA, BM

<u>Study supervision:</u> OA, BM

